# Diagnostic Accuracy of Large Language Models for Rare Diseases: A Systematic Review and Meta-Analysis

**DOI:** 10.64898/2026.03.26.26349194

**Authors:** Minh-Ha Nguyen, Chih-Ting Yang, Thomas A. Cassini, Fan Ma, Rizwan Hamid, Lisa Bastarache, Josh F. Peterson, Hua Xu, Lingyao Li, Siyuan Ma, Cathy Shyr

**Affiliations:** Department of Epidemiology, Vanderbilt University, Nashville, TN, USA; Department of Biostatistics, Vanderbilt University Medical Center, Nashville, TN, USA; Department of Pediatrics, Vanderbilt University Medical Center, Nashville, TN, USA; Department of Biomedical Informatics and Data Science, Yale School of Medicine, New Haven, CT, USA; Department of Biomedical Informatics, Vanderbilt University Medical Center, Nashville, TN, USA; School of Information, University of South Florida, Tampa, FL, USA

**Author notes:** Co-last and co-corresponding authors (Lingyao Li, Siyuan Ma, Cathy Shyr).

## Abstract

**Background:** Large language models (LLMs) have been evaluated as tools to assist rare disease diagnosis, yet evidence on their accuracy remains fragmented. We conducted a systematic review and meta-analysis to synthesize the available evidence on the diagnostic performance of LLMs, identify sources of heterogeneity, and evaluate the current evidence base for clinical translation.

**Methods:** We searched PubMed, Embase, Web of Science, Cochrane Library, arXiv, and medRxiv (January 2020–February 2026). Full-text articles and preprints were considered for inclusion. Eligible studies applied LLM-based systems to generate differential diagnoses for rare diseases and provided Recall@1 (R@1; proportion with the correct diagnosis ranked first). We pooled R@1 using Freeman–Tukey double arcsine transformation with DerSimonian–Laird random-effects models. Pre-specified subgroup analyses examined LLM knowledge augmentation strategy and input modality. Because both retained high residual heterogeneity, we conducted a post-hoc exploratory analysis of evaluation benchmark disease composition, mapping diseases from major benchmarks to Orphanet prevalence classifications. Risk of bias was assessed using a modified QUADAS-3 instrument.

**Findings:** We identified 902 records, of which 564 were screened and 15 studies were eligible. These 15 studies contributed 19 system–dataset entries to the meta-analysis (total N=39,529 cases). The pooled R@1 was 43.3% (95% CI 35.1–51.6; *I*^2^=99.6%). Augmented LLM systems (agent-based reasoning, retrieval, or fine-tuning; k=8) achieved R@1 of 52.5% (42.0–62.9) versus 35.4% (30.6–40.4) for standalone LLMs (k=11; p=0.004). Post-hoc exploratory analysis indicated that evaluation benchmark disease composition was associated with differences in diagnostic performance: R@1 was lower on the Phenopacket Store dataset, which contained a higher proportion of ultra-rare diseases (52.8%; k=2), than on RareBench (29.3%; k=6) at 21.7% (18.2–25.5) versus 52.0% (40.7–63.2; p<0.001). All 19 system-dataset entries were assessed to be at high risk of bias, most commonly due to potential data leakage and limited reproducibility. No study provided prospective clinical validation.

**Interpretation:** Diagnostic performance of LLM-based systems for rare diseases varied substantially across evaluation benchmarks. Post-hoc exploratory analysis indicated that performance was associated with benchmark disease composition. Performance was higher in benchmarks containing fewer ultra-rare diseases and in systems incorporating external knowledge at inference time. However, all included studies were at high risk of bias, and none reported prospective clinical validation. These findings highlight the need for prevalence-stratified evaluation benchmarks and independent prospective studies before clinical deployment.

**Funding:** This work was supported in part by the National Institutes of Health Common Fund, grant 15-HG-0130 from the National Human Genome Research Institute, U01NS134349 from the National Institute of Neurological Disorders and Stroke, R00LM014429 from the National Library of Medicine, and the Potocsnak Center for Undiagnosed and Rare Disorders.

## 1 Introduction

Rare diseases, each affecting fewer than one in 2,000 individuals, collectively affect over 300 million people worldwide [Nguengang Wakap et al., 2019]. Over 7,000 rare diseases have been described, with 70–80% having a genetic cause [Haendel et al., 2019]. Due to clinical heterogeneity and limited clinician familiarity, rare disease patients often undergo diagnostic odysseys characterised by repeated evaluations, unnecessary testing, and diagnostic delay averaging four to eight years [Haendel et al., 2019, Dawkins et al., 2018] . These delays have substantial medical, psychosocial, and economic consequences, including disease progression without appropriate treatment [Chung et al., 2022, Cohen and Biesecker, 2010, Tisdale et al., 2021, Carmichael et al., 2014] . Shortening the time to an accurate diagnosis is a major priority in rare disease care.

The reduced cost and increased availability of genome sequencing over the past decade have created new opportunities to improve rare disease diagnosis [Shyr and Liu, 2013], but these advances have shifted the diagnostic bottleneck towards knowledge synthesis and interpretation of complex clinical and genomic data [Splinter et al., 2018] . A typical exome or genome sequencing report can yield hundreds of candidate variants, and determining clinically relevance requires correlating subtle phenotypic patterns with thousands of possible rare diseases. This often exceeds individual clinician familiarity and is difficult to address using rule-based approaches. Computational phenotype-driven diagnostic support tools such as Exomiser [Smedley et al., 2015], Phen2Gene [Zhao et al., 2020], and LIRICAL [Robinson et al., 2020] were developed to address this challenge by systematically linking patient phenotypes to candidate diseases or genes [Robinson et al., 2008] . These tools can support diagnostic workflows in practice but rely on structured Human Phenotype Ontology (HPO) annotations that depend on accurate and comprehensive phenotype curation, making the process labour-intensive and difficult to scale.

To this end, recent advances in large language models (LLMs) offer innovative solutions to address these limitations. First, LLMs are trained on large-scale natural language corpora that can include biomedical sources such as medical literature, case reports, and gene-phenotype databases. This training may enable them to capture associations relevant to rare disease diagnosis without requiring explicitly engineered rule-based systems. [Shyr et al., 2025, Kanjee et al., 2023] . Second, unlike traditional HPO-based tools that primarily rely on structured phenotypes, LLMs can operate directly on unstructured clinical narratives, enabling them to interpret phenotypic descriptions and generate diagnostic hypotheses at scale and without manual curation. In practice, such systems could potentially function as clinical decision support tools that assist clinicians in generating differential diagnoses from phenotypic descriptions, prioritising candidate genetic variants, or guiding downstream genomic testing [Kim et al., 2024, Schaaf et al., 2020] . Since 2023, a growing body of studies has evaluated LLM-based approaches ranging from standalone prompting to retrieval-augmented systems and multi-agent frameworks designed to integrate phenotypic descriptions with genetic information [Shah, 2024, Lee et al., 2025, Zhou et al., 2025, Yang et al., 2025a] .

However, the reliability and clinical safety of LLM-generated differential diagnoses remain uncertain. Generative models can produce inaccurate or fabricated outputs [Xu et al., 2024, Ahmad et al., 2023], and their performance can vary depending on prompt strategy, training data, and access to external knowledge sources [Schaaf et al., 2020, Zhuo et al., 2024] . Reported diagnostic performance varies substantially across studies, reflecting differences in evaluation benchmarks, input modalities, LLM augmentation strategies, and evaluation protocols. In addition, many evaluations rely on curated cases for benchmarks [Reese et al., 2026, Chimirri et al., 2025], which may not reflect the distribution and information available on diseases encountered in real clinical practice. As a result, the current evidence base remains fragmented and difficult to interpret.

Despite the rapidly increasing interest in LLM-based diagnostic tools, their diagnostic accuracy and sources of performance variability have not been systematically synthesized across studies. Understanding the conditions under which these systems perform well is essential to evaluate their potential role in clinical decision support. To address this gap, we conducted a systematic review and meta-analysis to: **(1) quantify the pooled diagnostic accuracy of LLM-based systems for rare diseases; (2) identify the major sources of heterogeneity in diagnostic performance across studies; and (3) assess the methodological quality and clinical readiness of the current evidence base**.

## 2 Methods

### 2.1 Protocol and Registration

This systematic review and meta-analysis followed PRISMA-DTA guidelines [McInnes et al., 2018] . The study protocol was registered prospectively on the Open Science Framework [Nguyen et al., 2026] before data extraction.

### 2.2 Search Strategy

We searched six databases — PubMed/MEDLINE, Embase (Ovid), Web of Science Core Collection, Cochrane Library, arXiv, and medRxiv — from January 2020 to February 2026. The search strategy combined terms for large language models and artificial intelligence with terms for rare diseases, using both free-text and controlled vocabulary (MeSH/Emtree) where applicable. The strategy was iteratively refined to balance sensitivity and specificity against a validation set of known eligible studies (Supplementary Appendix). Complete search queries are provided in the Supplementary Appendix. No language restrictions were applied. Full-text articles and preprints were considered for inclusion.

### 2.3 Eligibility Criteria

Studies were eligible if they: (1) evaluated a system using at least one LLM as the primary diagnostic reasoning component; (2) assessed diagnosis of rare diseases using a defined evaluation cohort of more than 10 cases; and (3) reported strict top-1 diagnostic accuracy (R@1). We excluded studies reporting only Recall@k (*k* > 1) or “mentioned in differential” outcomes without extractable R@1, studies evaluating gene prioritisation rather than disease diagnosis, studies where the LLM served only as a component within a non-LLM ranking algorithm, review articles, or editorials.

### 2.4 Screening and Data Extraction

All stages of screening, full-text assessment, data extraction, and quality assessment were done independently by at least two authors. Two reviewers independently screened titles and abstracts (MHN and one of CS, SM, LL, or CY). Inter-rater agreement for title and abstract screening was 92.0% (Cohen’s *κ*=0.64). Full-text assessment was performed against pre-specified PIRD criteria; with inter-rater agreement of 76.9% (*κ*=0.54) by two independent authors (MHN and one of CS, SM, LL, or CY). Disagreements over inclusion or data extraction were resolved by consensus.

For each included study, we extracted study characteristics (authors, year, publication type), system characteristics (architecture, base LLM, augmentation strategy), evaluation benchmark dataset, sample size, input modality (structured HPO terms, unstructured clinical text, or combined phenotype and genetic data), and R@1 with numerator and denominator.

### 2.5 Meta-Analysis Entry Selection

To minimise within-paper correlation, we applied pre-specified entry selection rules registered in the study protocol: (1) extract the numbers reported in the abstract of the manuscript, where multiple numbers were reported, we gave priority to results on publicly available benchmarks over institutional data; (2) include a maximum of one baseline plus one augmented entry per paper on the same benchmark; (3) include only strict R@1; and (4) require *N* > 10.

### 2.6 Risk of Bias Assessment

Risk of bias was assessed using a modified QUADAS-3 [Tomlinson et al., 2025] instrument adapted for LLM diagnostic evaluation studies, comprising seven signalling domains: participant selection (D1), index test conduct (D2), target condition definition (D3), flow and timing (D4), data leakage (D5), reproducibility (D6), and evaluation fairness (D7). The adapted instrument is provided in the Supplementary Appendix. Assessments were performed by one reviewer (MHN) and independently verified by a second reviewer (one of CS, SM, LL, or CY), with disagreements resolved by discussion. An overall judgment of high risk of bias was assigned if any domain was rated high. The rationales are given in Supplementary Table S5.

### 2.7 Benchmark Disease Prevalence Analysis

To investigate whether benchmark disease composition contributed to performance heterogeneity, we mapped diseases from four commonly used evaluation benchmarks to Orphanet prevalence classifications (Product 9, epidemiology data). For the Phenopacket Store dataset (694 diseases), we extracted OMIM identifiers from GA4GH phenopacket files [Jacobsen et al., 2022] and mapped to Orphanet via OMIM-to-OrphaCode mapping (mapping rate: 49.1%). For RareBench (301 diseases), MAC Medline (301 diseases), and the Chinese Rare Disease List (66 diseases), disease names were mapped to Orphanet via programmatic search and manual curation (mapping rates: 88–91%). Diseases were classified according to Orphanet prevalence categories as ultra-rare (<1 per million), rare (1–9 per million), or higher-prevalence (≥1 per 100,000). The GA4GH Phenopacket format embeds pre-identified causal variants (mean 1.1 per case) with the American College of Medical Genetics (ACMG) pathogenicity classifications [Richards et al., 2015] ; this was verified by direct inspection of 9,588 phenopacket files (Supplementary Methods).

### 2.8 Statistical Analysis

The primary outcome was Recall@1 (R@1), defined as the proportion of cases in which the correct diagnosis was ranked first in the generated differential diagnosis. Effect sizes were computed using the Freeman–Tukey double arcsine transformation for proportions. Pooled estimates were calculated using the DerSimonian–Laird random-effects model with back-transformation using the harmonic mean of sample sizes. Heterogeneity was assessed using *I*^2^ and *τ*^2^. Differences between subgroups were tested using the *Q*_between_ statistic. Pre-specified subgroup analyses examined diagnostic performance by system architecture (augmented versus baseline) and input modality (structured HPO versus unstructured clinical text). Because both pre-specified subgroups retained high residual heterogeneity (*I*^2^ > 95%), we conducted a post-hoc exploratory analysis of benchmark family (RareBench versus Phenopacket Store) as a candidate moderator, followed by Orphanet prevalence mapping of benchmark disease compositions. Sensitivity analyses included leave-one-out analyses, exclusion of Phenopacket Store entries (to account for shared case pools), and exclusion of small datasets (*N* < 100). All analyses were conducted in R (metafor).

## 3 Results

### Study Selection and Characteristics

We identified 902 records across six databases (PubMed n=183, Embase n=409, Web of Science n=221, Cochrane n=6, arXiv n=68, medRxiv n=15). After removing 338 duplicates, 564 unique records were screened by title and abstract, of which 78 underwent full-text assessment (Figure 1). After full-text exclusions for non-R@1 metrics, gene prioritisation tasks, absence of an LLM as the primary diagnostic reasoning component, or insufficient sample size, 15 studies met all inclusion criteria and contributed 19 system-dataset entries to the quantitative meta-analysis (Table 1) [Reese et al., 2026, Chimirri et al., 2025, Yang et al., 2025b, Zhao et al., 2026, Wang et al., 2026, Zheng et al., 2025, Chen et al., 2024a, 2025, Rose et al., 2025, Schumacher et al., 2025, Chen et al., 2024b, AlDin et al., 2026, Ilić et al., 2025, Ao et al., 2025, do Olmo et al., 2024] . Four papers contributed matched baseline–augmented system comparisons evaluated on the same benchmark; the remaining eleven contributed single entries. Studies were published between 2024 and 2026, with 17 of 19 entries from 2025 or later. Base LLMs included GPT-4/4o (11 entries), Llama variants (3), Claude 3 Opus/3.5 Sonnet (2), and other models including o1-preview, DeepSeek-V3, and MedGemma-27B. Evaluation benchmarks included RareBench sub-benchmarks, the Phenopacket Store, MIMIC-IV - Rare, and institution-specific datasets.

**Figure 1.**
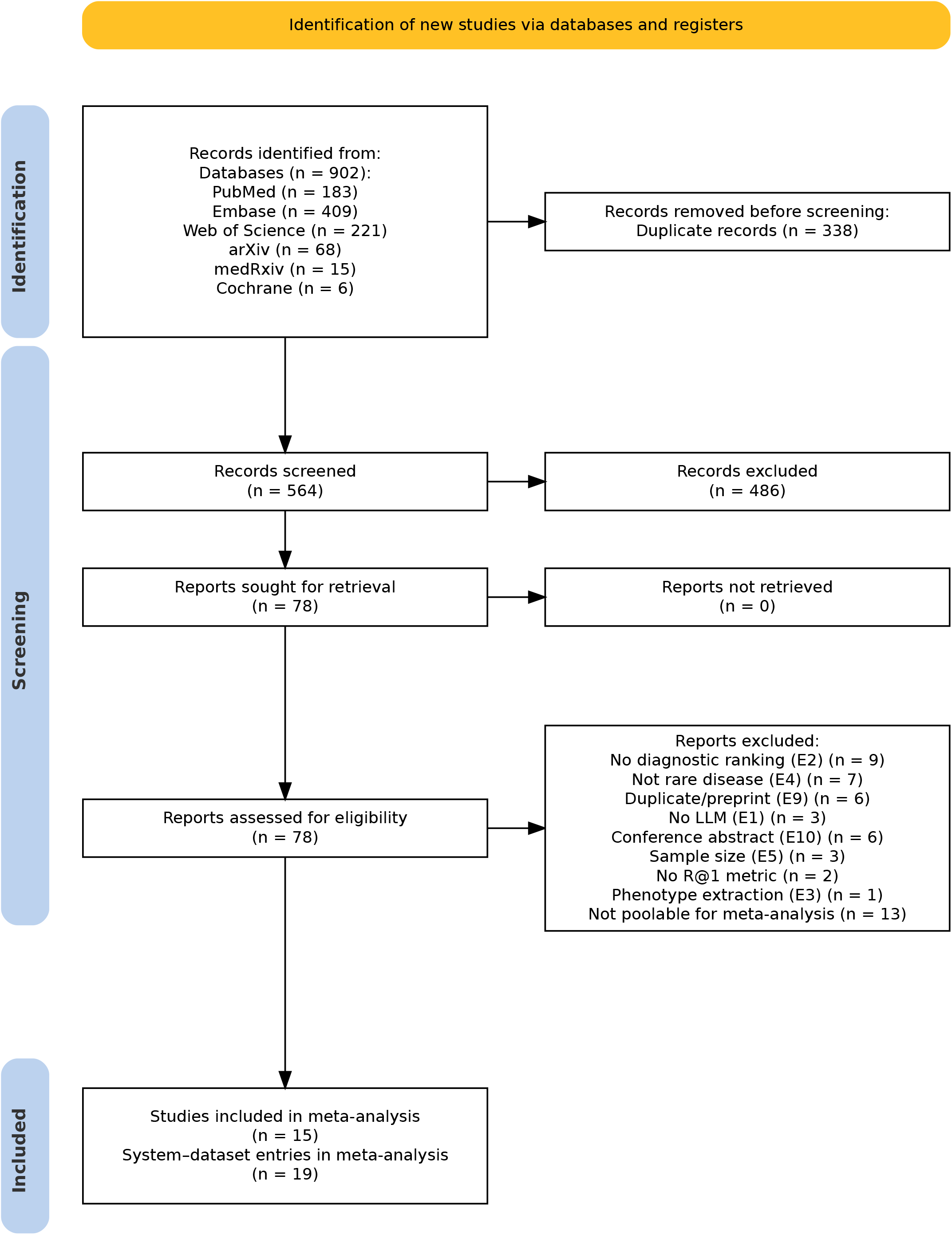
PRISMA 2020 flow diagram showing identification, screening, and inclusion of studies. Of 902 records identified across six databases, 564 unique records were screened, 78 assessed at full text, and 15 studies (19 system–dataset entries) included in the meta-analysis.

**Table 1:**
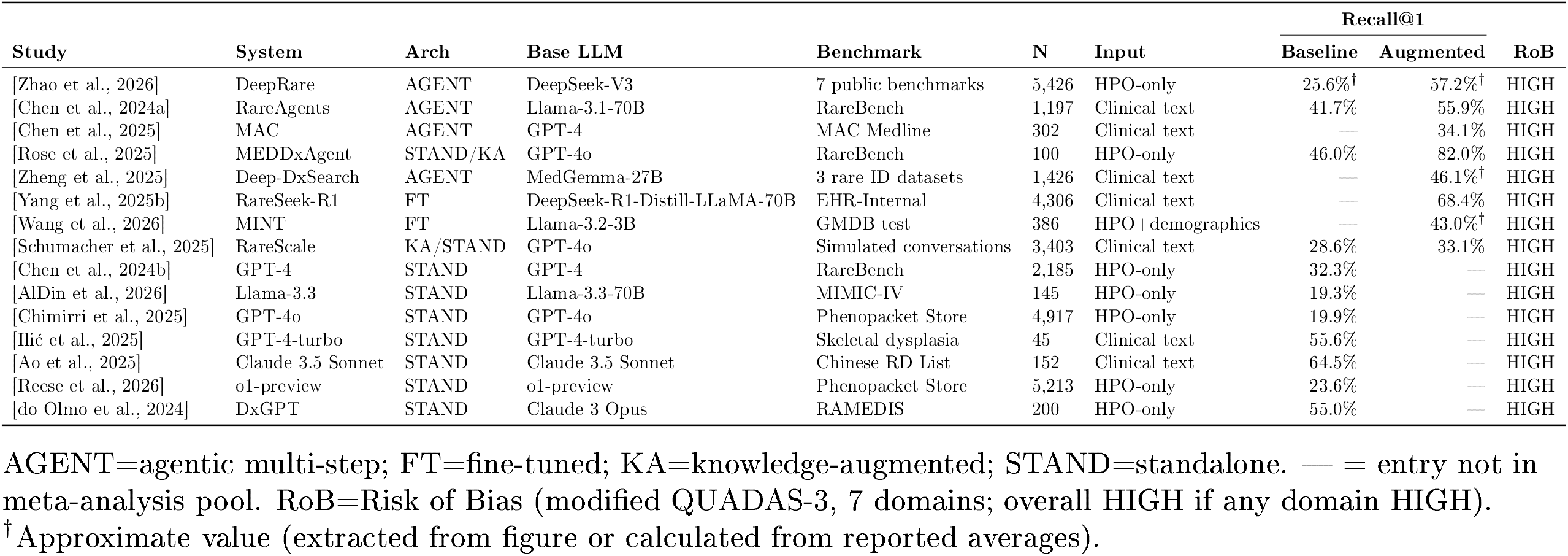
Characteristics of the 15 studies (19 system–dataset entries) included in the meta-analysis. Sorted by R@1 within augmentation groups.

### Overall Diagnostic Accuracy

The pooled R 1 across 19 system–dataset entries was 43.3% (95% CI 35.1–51.6; *I*^2^=99.6%, *τ*^2^=0.134; Figure 2). The high heterogeneity indicates substantial variation in diagnostic performance across evaluation settings. Leave-one-out sensitivity analysis showed the pooled R@1 estimate was stable (range 41.1–44.7%). Excluding two Phenopacket Store entries with overlapping case pools increased the pooled R@1 to 46.0% (37.7–54.5%).

**Figure 2.**
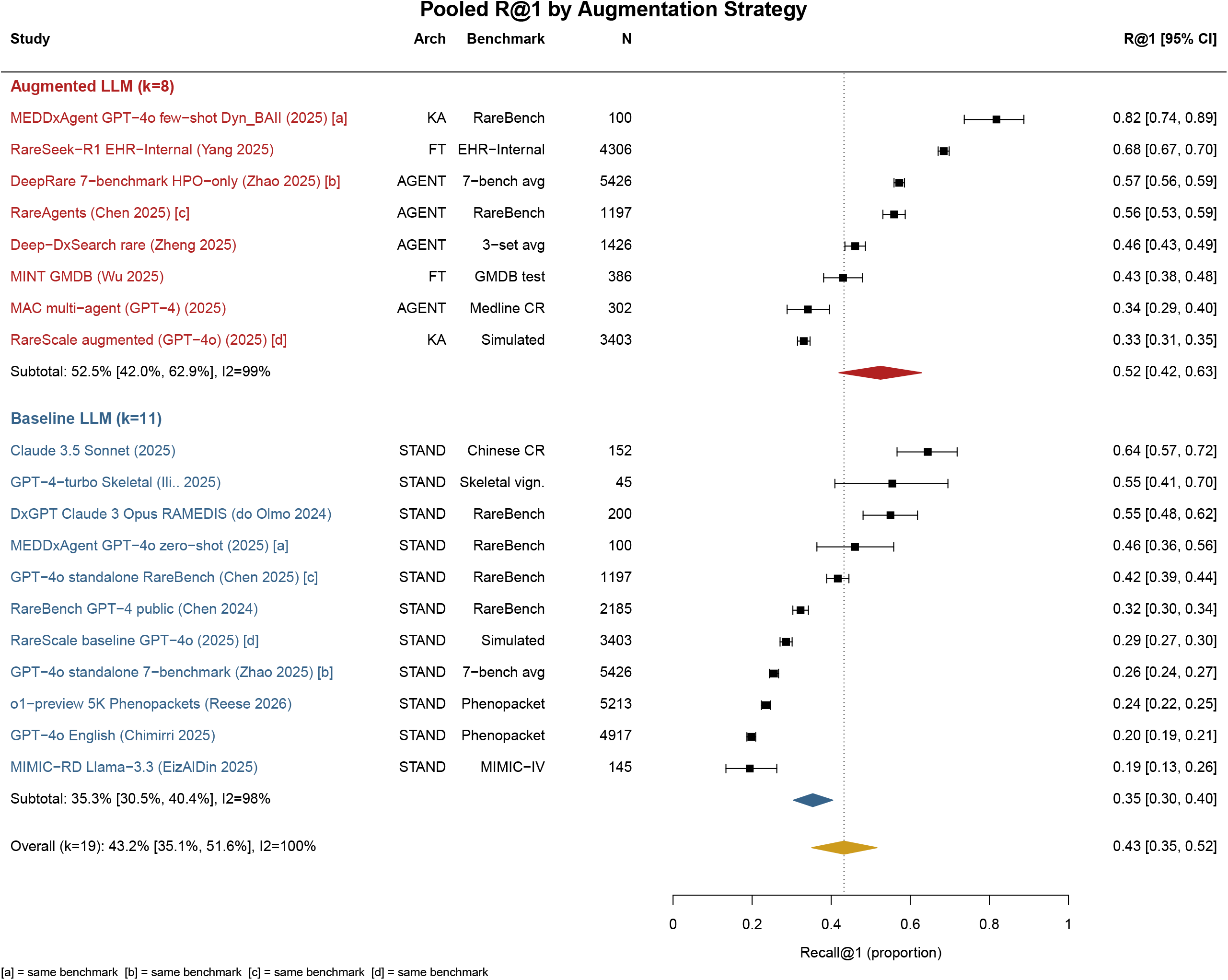
Forest plot of pooled Recall@1 (R@1) by augmentation strategy. Augmented systems (k=8; agent-based, retrieval-augmented, or fine-tuned) achieved higher pooled R@1 (52.5%, 95% CI 42.0–62.9) than standalone LLMs (k=11; 35.4%, 30.6–40.4; p=0.004). Overall pooled R@1 was 43.3% (35.1–51.6; *I*^2^=99.6%).

### Benchmark Composition and Diagnostic Performance

Pre-specified subgroup analyses (augmentation strategy, input modality) did not resolve the high heterogeneity observed in the overall pool (*I*^2^ > 95% within both subgroups). We therefore examined benchmark family as a post-hoc exploratory moderator. Diagnostic performance differed substantially across evaluation benchmarks. In the meta-analysis pool, RareBench entries (k=6) yielded a pooled R@1 of 52.0% (40.7–63.2), compared with 21.7% (18.2–25.5; *I*^2^=95%) on the Phenopacket Store (k=2; *Q*_between_–26.53, p<0.001). An expanded comparison including all available R@1 values from included studies on these two benchmarks (RareBench k=8, Phenopacket Store k=6; Figure 3, panel A) illustrates this pattern. On the Phenopacket Store, four independent entries from four studies using standalone or fine-tuned models had R@1 in the 19.2–23.6% range, while a medical-domain fine-tuned model (Meditron3-70B, 15.4%) and a knowledge-graph augmented system (RareSeek+GraphRAG [Yang et al., 2025b] 41.4%) bracketed this range. On RareBench, R@1 ranged from 14.6% (Gemini Pro, standalone) to 65.8% (DeepRare [Zhao et al., 2026] agent-based), with higher performance observed for systems incorporating augmentation within the same benchmark.

**Figure 3.**
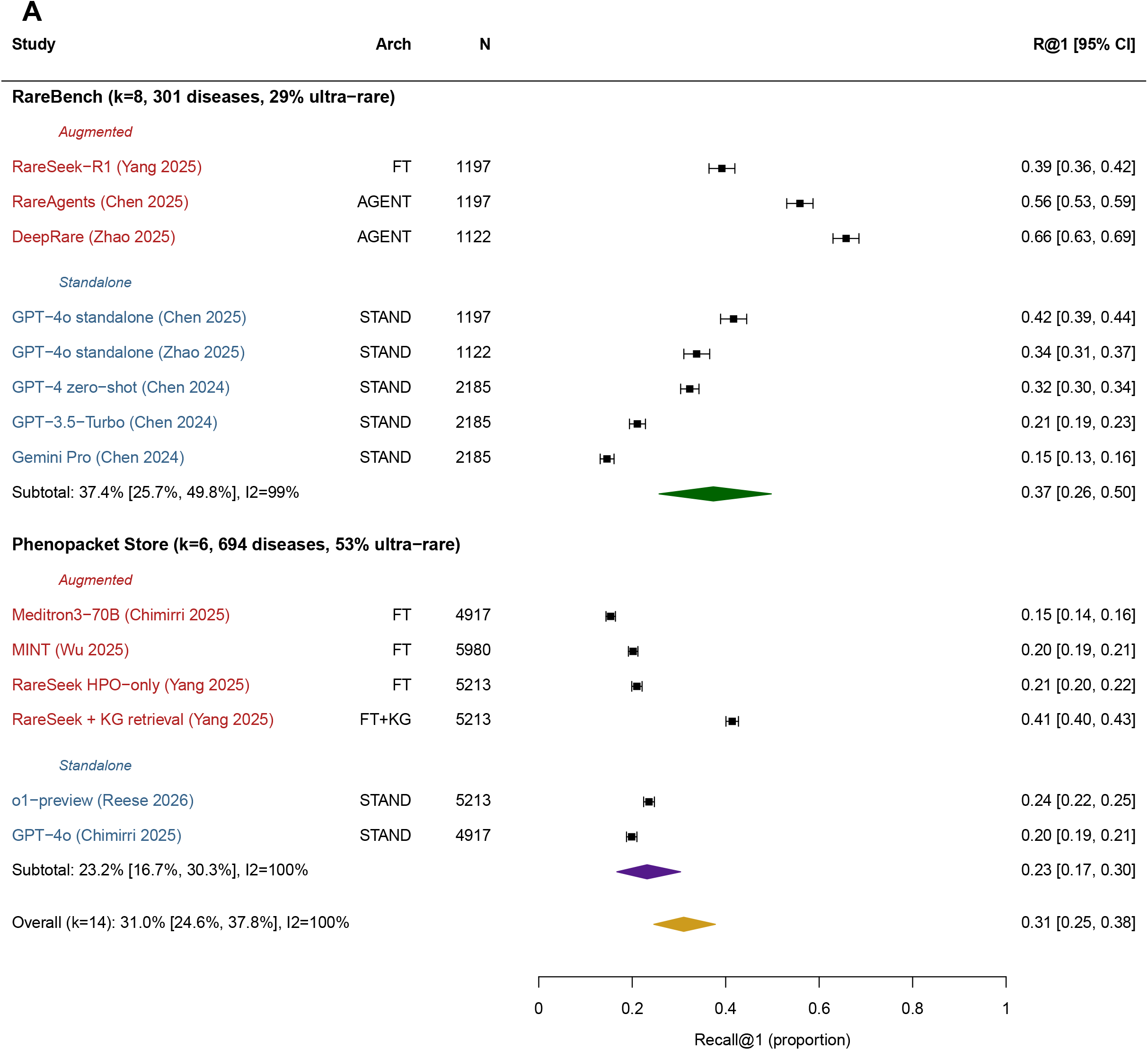
Illustrative forest plot and prevalence-accuracy relationship from LLM systems (post-hoc exploratory analysis). **Panel :A** Expanded forest plot comparing R@1 across RareBench (k=8) and Phenopacket Store (k=6) benchmarks. RareBench R@1 ranged from 14.6% to 65.8%; Phenopacket Store from 15.4% to 41.4%.

**Figure 3.**
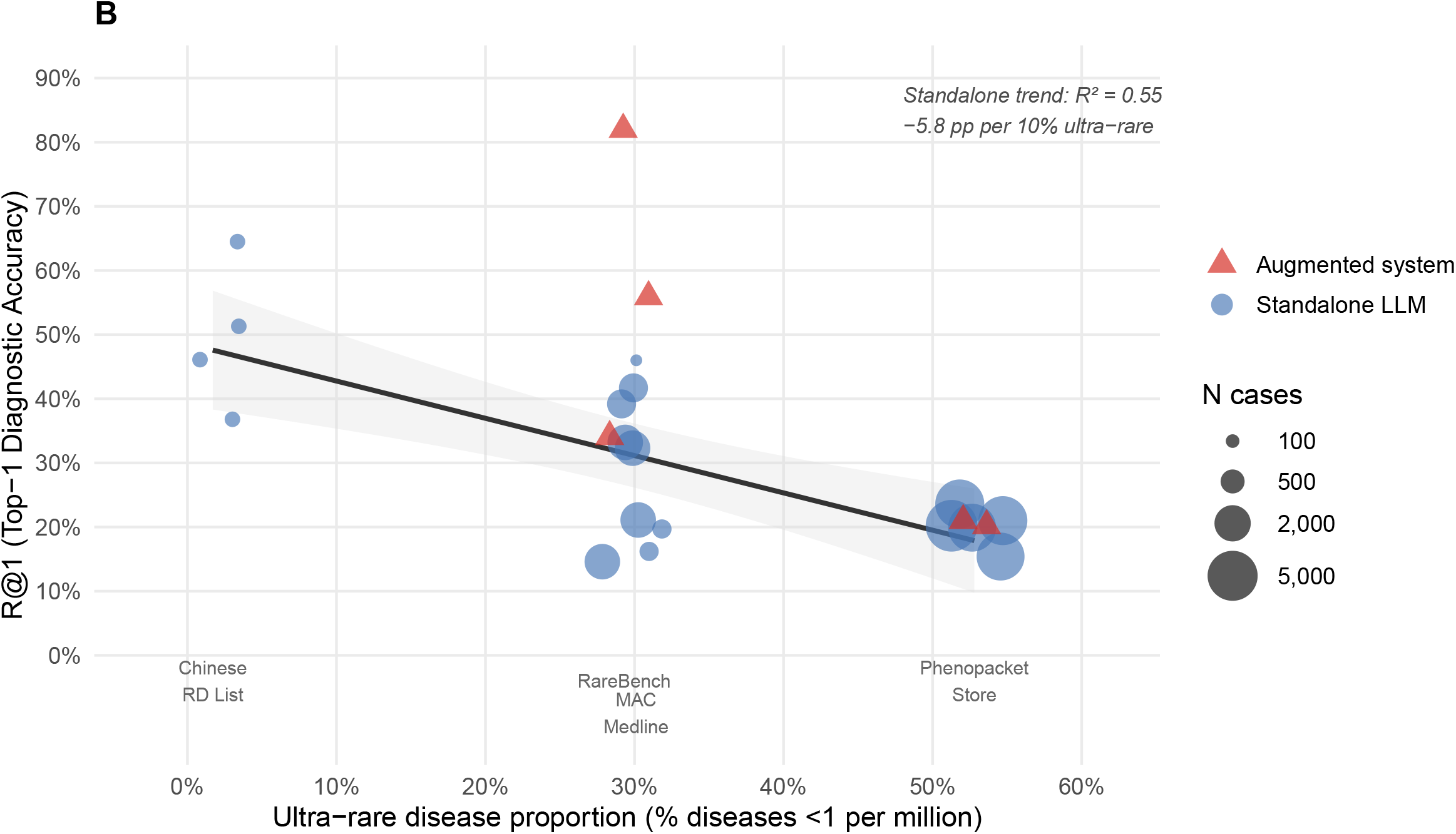
**Panel B:** Relationship between benchmark ultra-rare disease proportion and standalone LLM diagnostic accuracy. The negative trend (R^2^=0.55, −5.8 percentage points per 10% increase in ultra-rare proportion) suggests that benchmarks with higher proportions of ultra-rare diseases yield lower R@1. Circle size proportional to sample size.

Orphanet prevalence mapping of benchmark disease compositions showed substantial differences across datasets (Table 2; Figure 3, panel B). The Phenopacket Store comprises 52.8% ultra-rare diseases (<1 per million), compared with 29.3% for RareBench and 1.7% for the Chinese Rare Disease List. Across included studies, best-performing standalone LLMs achieved R@1 values of 19.9–23.6% on the Phenopacket Store, 32.3–46.% on RareBench, and 64.5% on the Chinese Rare Disease List. Within the same prevalence range, performance also varied across benchmarks. For example, MAC Medline and RareBench have similar ultra-rare proportions (30.1% vs 29.3%) but different R@1 ranges (16–20% vs 32–46%). Differences in dataset structure were also observed. RareBench’s sub-benchmarks (e.g., RAMEDIS with 624 cases across 74 diseases) included multiple cases per disease, whereas MAC Medline comprised 301 unique diseases drawn from individual case reports (1 case per disease). Within-study comparisons further showed variability across benchmarks. In the DeepRare [Zhao et al., 2026] evaluation across seven public sub-benchmarks, the same system’s R@1 ranged from 29% on MIMIC-IV -Rare (355 diseases) to 73% on RAMEDIS. Similarly, MME and LIRICAL, which have comparable median disease prevalence (0.5 per million), yielded different R@1 values (70% vs 56% for DeepRare) (Table S7).

**Table 2:** Benchmark characteristics and disease prevalence composition. Ordered by ascending proportion of ultra-rare diseases. Prevalence mapped via Orphanet Product 9 epidemiology data.

| Benchmark | Source | N cases | N diseases | Cases/dis | Input | % ultra-rare | Standalone R@1 | Augmented R@1 | k |
| --- | --- | --- | --- | --- | --- | --- | --- | --- | --- |
| Chinese RD List | Chinese case repos. | 152 | 66 | 2.3 | Text | 1.7% | 64.5% | — | 1 |
| RareBench-RAMEDIS* | Scientist-uploaded | 624 | 74 | 8.4 | HPO | ~5% | 35.5–42.5% | 72.6% (DeepRare) | 1 |
| RareBench (agg.)* | Multi-source (public) | 1,122–2,185 | 301–421 | 3.7 | Mixed | 29.3% | 14.6–46.0% | 65.8% (DeepRare) | 5 |
| MAC Medline | Case reports (post-2022) | 302 | 301 | 1.0 | Text | 30.1% | 16.2–19.7% | 34.1% (MAC) | 1 |
| RareBench-LIRICAL* | Literature-derived | 370 | 252 | 1.5 | HPO | ~45% | 25.7–38.1% | 56.0% (DeepRare) | 1 |
| Phenopacket Store | GA4GH phenopackets | 4,917–5,980 | 360–694 | 7.2–8.5 | HPO | 52.8% | 15.4–23.6% | 41.4% (RareSeek+KG) | 4 |

Mapping rates: PS 49.1%, RareBench 90.7%, MAC 88.4%, Chinese RD 90.9%. \*RAMEDIS, LIRICAL are RareBench sub-benchmarks.

### LLM Augmentation Strategy and Generalizability

Systems incorporating any form of inference-time augmentation — agent-based multi-step reasoning, retrieval augmentation, or domain-specific fine-tuning (k=8) — achieved higher pooled R@1 than standalone LLM prompting approaches (k=11). The pooled R@1 was 52.5% (42.0–62.9) for augmented systems compared with 35.4% (30.6–40.4) for standalone LLMs with standard prompting (k=11; p=0.004; Figure 2). Four papers provided matched within-paper comparisons on the same benchmark. In all four papers, augmented systems achieved higher R@1 than their baseline counterparts, with improvements ranging from 4.5 to 36 percentage points (Figure S1).

Two fine-tuned systems (MINT [Wang et al., 2026], RareSeek [Yang et al., 2025b]) contributed entries to the augmented pool. In both cases, diagnostic accuracy differed between evaluation settings. For MINT, external validation had R@1 of 47.6% on diseases present in the training set compared to 7.0% on diseases not included in the training set. On unseen diseases, a retrieval-augmented baseline (13.8%) outperformed the fine-tuned model (7.0%). Similarly, RareSeek achieved 68.4% R@1 on institutional data used for fine-tuning and 39.2% on an independent benchmark (RareBench).

### Input Modality

Input modality (structured HPO terms versus unstructured clinical text) was not a significant moderator in the pooled analysis (39.6% versus 47.3%; p=0.35). Only two included studies incorporated genetic variant data. In one study, inclusion of exome sequencing data yielded higher R@1 compared with HPO-only input (R@1 of 69.1% vs 57.2%; DeepRare [Zhao et al., 2026]). In the second study, variant information was provided as pre-identified pathogenic variants within the GA4GH Phenopacket format, and variant-only input yielded R@1 of 47.9% (RareSeek [Yang et al., 2025b]). Because these approaches differed substantially in design and task definition, they were not included in the formal meta-analysis pool.

### Risk of Bias

All 19 system-dataset entries were rated to be at high risk of bias (Table 3). The most commonly identified concern was potential data leakage between evaluation datasets and LLM pre-training or fine-tuning corpora (18 of 19 entries). No study provided independent replication by an external group, prospective clinical validation, or evaluation of real-world clinical outcomes such as time to diagnosis.

**Table 3:** Risk of bias summary (modified QUADAS-3). H=HIGH, L=LOW risk of bias. Overall risk was rated HIGH if any domain was HIGH.

| Study | System | D1 | D2 | D3 | D4 | D5 | D6 | D7 | Overall |
| --- | --- | --- | --- | --- | --- | --- | --- | --- | --- |
| [Chimirri et al., 2025] | GPT-4o | L | L | L | L | H | H | L | H |
| [Ilić et al., 2025] | GPT-4-turbo | L | L | L | L | H | H | L | H |
| [Wang et al., 2026] | MINT | L | L | L | L | H | L | L | H |
| [Ao et al., 2025] | Claude 3.5 | L | L | L | L | H | H | L | H |
| [Chen et al., 2025] | MAC | L | L | L | L | L | H | L | H |
| [Reese et al., 2026] | o1-preview | L | L | L | L | H | L | L | H |
| [Chen et al., 2024b] | RareBench | L | L | L | L | H | H | L | H |
| [Chen et al., 2024a] | RareAgents | L | H | L | L | H | L | H | H |
| [Schumacher et al., 2025] | RareScale | L | L | H | L | H | H | L | H |
| [Rose et al., 2025] | MEDDxAgent | L | L | L | L | H | L | L | H |
| [Zhao et al., 2026] | DeepRare | L | L | L | L | H | L | L | H |
| [Zheng et al., 2025] | Deep-DxSearch | L | L | L | L | H | L | L | H |
| [Yang et al., 2025b] | RareSeek | H | L | H | L | H | H | L | H |
| [AlDin et al., 2026] | MIMIC-RD | L | L | H | L | H | ? | L | H |
| [do Olmo et al., 2024] | DxGPT | L | H | L | L | H | H | L | H |

D1=Participant selection; D2=Index test; D3=Target condition; D4=Flow/timing; D5=Data leakage; D6=Reproducibility; D7=Evaluation fairness. ?=Unclear.

## 4 Discussion

To our knowledge, this is the first systematic review and meta-analysis evaluating the diagnostic performance of LLM-based systems for rare disease diagnosis. Across 19 system–dataset evaluations from 15 studies, LLM-based systems yielded a pooled R@1 of 43.3% (95% CI 35.1–51.6), with high heterogeneity (*I*^2^ =99.6%). Several findings are notable: LLM-based systems processing free-text narratives performed comparably to those relying on structured HPO annotations, and incorporating genetic information alongside phenotypic input was associated with higher diagnostic accuracy. These observations suggest that multimodal inputs may be associated with improved diagnostic performance. Nevertheless, at this level of heterogeneity, the pooled estimates should be interpreted as a descriptive summary across diverse evaluation settings rather than a predictor of real-world performance. Overall, these findings suggest promising capabilities of LLMs to operate directly on unstructured clinical narrative and synthesize phenotypic and genetic information to support rare disease diagnosis. However, reported diagnostic accuracy varies substantially across studies and appears associated with characteristics of the evaluation benchmarks, underscoring the need for more standardised evaluation before clinical translation.

We highlight three observations from our study that are relevant for clinical translation of LLM-based systems. First, diagnostic performance varied substantially across evaluation benchmarks. **The 30 percentage-point difference between the benchmarks, RareBench (52.0%) and Phenopacket Store (21.7%), exceeds differences observed across model architectures, augmentation strategies, or input modalities**. Orphanet prevalence mapping showed that benchmarks differed substantially in the proportion of ultra-rare diseases, and performance tended to be lower in datasets with a higher proportion of such conditions (Table S1). Within similar prevalence ranges, diagnostic performance also varied across datasets with different structures and disease representation. For example, benchmarks containing multiple cases per disease or well-characterised conditions showed higher R@1 than those of single-case reports spanning many distinct diseases. These patterns suggest that differences in benchmark composition, including disease prevalence, representation, and available clinical information, may contribute to the observed heterogeneity in performance. **These findings have implications for interpreting reported accuracy in the literature. High performance on public benchmark datasets might not generalize across broader rare disease populations, especially those enriched for ultra-rare or sparsely described conditions. Reporting benchmark composition, including disease prevalence, will be important for contextualizing evaluation results**.

Second, systems incorporating inference-time augmentation strategies, including retrieval, agent-based reasoning, or fine-tuning, were associated with higher diagnostic accuracy than standalone LLMs (52.5% vs. 35.4%). Agent-based architectures that combine retrieval with iterative hypothesis testing showed the largest and most consistent gains. On the Phenopacket Store benchmark, where standalone and fine-tuned models yielded R@1 ranging between 19 and 24%, knowledge-graph retrieval from Orphanet and OMIM [Yang et al., 2025b] increased R@1 to 41.4%, demonstrating that providing structured phenotype-disease evidence at inference improves accuracy. Approaches that incorporate external knowledge sources at inference time, such as retrieval from curated knowledge bases, offer a plausible strategy for supporting diagnosis in settings where relevant disease information is sparse or evolving. Whether these approaches translate into improved clinical outcomes, however, remains to be established.

Third, the current evidence base does not support clinical deployment. All included studies were retrospective, and all risk-of-bias assessments were rated high, with common concerns including potential data leakage and lack of independent validation. No study evaluated prospective clinical use or clinically meaningful endpoints such as impact on time to diagnosis or real-world patient outcomes. One study reported improved physician diagnostic accuracy when using an LLM-based system as decision support [Zheng et al., 2025], but such evidence remains limited. The included studies primarily focused on performance on curated benchmark datasets rather than real-world clinical datasets, where information is often incomplete and diagnostic decisions are made under time and resource constraints.

Several limitations should be considered. First, the DerSimonian–Laird model does not account for within-paper correlation, although we applied pre-defined selection rules to mitigate this effect. Second, substantial heterogeneity across studies reflects differences in benchmarks, input modalities, evaluation protocols, and disease composition, limiting comparability of pooled estimates. Third, some studies used constrained candidate lists rather than open-ended diagnosis generation, which should be considered when interpreting R@1 across studies. Fourth, the benchmark prevalence analysis was based on a subset of benchmarks with variable Orphanet mapping rates (49–91%); estimates of ultra-rare disease representation are therefore approximate. Finally, four entries relied on approximate values extracted from figures. In addition, our review was scoped to rare disease diagnosis; studies focusing on LLM-based gene prioritisation [Lee et al., 2025, Zhou et al., 2025, Neeley et al., 2024] and hybrid LLM-traditional systems [Yang et al., 2025a, Qi et al., 2026] were assessed but excluded from quantitative synthesis as they address complementary clinical tasks.

These findings highlight several considerations for future studies. First, studies should report performance stratified by disease prevalence or equivalent measures of rarity to enable meaningful comparison across benchmarks. Second, evaluation should include disease-level holdout sets to better assess generalizability to previously unseen conditions. Third, routine assessment of potential data leakage or contamination between training and evaluation data will be critical for interpreting performance. Fourth, prospective studies are needed to evaluate clinical utility, including effects on diagnostic accuracy, time to diagnosis, and clinical decision-making. Given the rapid growth of rare disease knowledge, LLM-based approaches that integrate up-to-date external information sources might offer advantages in incorporating newly available information to assist clinicians in diagnosis, but their impact in real-world settings remains to be established.

## Supporting information

PRISMA-DTA-checklist

## Data Availability

Meta-analysis data, computation scripts, forest plot code, and benchmark prevalence mapping files will be made available at a public GitHub repository upon publication of the manuscript.

