## Supplementary material for "Diagnostic Accuracy of Large Language Models for Rare Diseases: A Systematic Review and Meta-Analysis": PRISMA-DTA-checklist

| **Section/topic** | **#** | **PRISMA-DTA Checklist Item** | **Reported on page #** |
| --- | --- | --- | --- |
| **TITLE / ABSTRACT** | | |  |
| Title | 1 | Identify the report as a systematic review (+/- meta-analysis) of diagnostic test accuracy (DTA) studies. | Title page (p. 1) |
| Abstract | 2 | Abstract: See PRISMA-DTA for abstracts. | Abstract (pp. 1-2) |
| **INTRODUCTION** | | |  |
| Rationale | 3 | Describe the rationale for the review in the context of what is already known. | Introduction (pp. 2-3) |
| Clinical role of index test | D1 | State the scientific and clinical background, including the intended use and clinical role of the index test, and if applicable, the rationale for minimally acceptable test accuracy (or minimum difference in accuracy for comparative design). | Introduction, paragraphs 2-3 (pp. 2-3); Methods: Eligibility Criteria (p. 4) |
| Objectives | 4 | Provide an explicit statement of question(s) being addressed in terms of participants, index test(s), and target condition(s). | Introduction, paragraph 4 -- three objectives (p. 3) |
| **METHODS** | | |  |
| Protocol and registration | 5 | Indicate if a review protocol exists, if and where it can be accessed (e.g., Web address), and, if available, provide registration information including registration number. | Methods: Protocol and Registration (p. 4); OSF osf.io/zebn5; DOI: 10.17605/OSF.IO/FMABN |
| Eligibility criteria | 6 | Specify study characteristics (participants, setting, index test(s), reference standard(s), target condition(s), and study design) and report characteristics (e.g., years considered, language, publication status) used as criteria for eligibility, giving rationale. | Methods: Eligibility Criteria (p. 4) |
| Information sources | 7 | Describe all information sources (e.g., databases with dates of coverage, contact with study authors to identify additional studies) in the search and date last searched. | Methods: Search Strategy (p. 3); Supplementary Appendix |
| Search | 8 | Present full search strategies for all electronic databases and other sources searched, including any limits used, such that they could be repeated. | Supplementary Appendix (full search strategies) |
| Study selection | 9 | State the process for selecting studies (i.e., screening, eligibility, included in systematic review, and, if applicable, included in the meta-analysis). | Methods: Screening and Data Extraction (p. 4); Figure 1 (PRISMA flow, p. 7) |
| Data collection process | 10 | Describe method of data extraction from reports (e.g., piloted forms, independently, in duplicate) and any processes for obtaining and confirming data from investigators. | Methods: Screening and Data Extraction (p. 4) |
| Definitions for data extraction | 11 | Provide definitions used in data extraction and classifications of target condition(s), index test(s), reference standard(s) and other characteristics (e.g. study design, clinical setting). | Methods: Eligibility Criteria (p. 4); Meta-Analysis Entry Selection (p. 4) |
| Risk of bias and applicability | 12 | Describe methods used for assessing risk of bias in individual studies and concerns regarding the applicability to the review question. | Methods: Risk of Bias Assessment (p. 5); Supplementary Appendix (modified QUADAS-3 instrument) |
| Diagnostic accuracy measures | 13 | State the principal diagnostic accuracy measure(s) reported (e.g. sensitivity, specificity) and state the unit of assessment (e.g. per-patient, per-lesion). | Methods: Statistical Analysis (p. 5) -- R@1 as primary measure, per-case unit |
| Synthesis of results | 14 | Describe methods of handling data, combining results of studies and describing variability between studies. This could include, but is not limited to: a) handling of multiple definitions of target condition. b) handling of multiple thresholds of test positivity, c) handling multiple index test readers, d) handling of indeterminate test results, e) grouping and comparing tests, f) handling of different reference standards | Methods: Statistical Analysis (p. 5-6); Meta-Analysis Entry Selection (p. 5) |

Page 1 of 2

| **Section/topic** | **#** | **PRISMA-DTA Checklist Item** | **Reported on page #** |
| --- | --- | --- | --- |
| Meta-analysis | D2 | Report the statistical methods used for meta-analyses, if performed. | Methods: Statistical Analysis (p. 5) -- Freeman-Tukey double arcsine + DerSimonian-Laird random-effects |
| Additional analyses | 16 | Describe methods of additional analyses (e.g., sensitivity or subgroup analyses, meta-regression), if done, indicating which were pre-specified. | Methods: Statistical Analysis (p. 5) -- pre-specified subgroup analyses (augmentation, input modality); post-hoc exploratory benchmark analysis; sensitivity analyses |
| **RESULTS** | | |  |
| Study selection | 17 | Provide numbers of studies screened, assessed for eligibility, included in the review (and included in meta-analysis, if applicable) with reasons for exclusions at each stage, ideally with a flow diagram. | Results: Study Selection and Characteristics (p. 6); Figure 1 |
| Study characteristics | 18 | For each included study provide citations and present key characteristics including: a) participant characteristics (presentation, prior testing), b) clinical setting, c) study design, d) target condition definition, e) index test, f) reference standard, g) sample size, h) funding sources | Results: Study Selection and Characteristics (p. 6); Table 1 |
| Risk of bias and applicability | 19 | Present evaluation of risk of bias and concerns regarding applicability for each study. | Results: Risk of Bias (p. 8); Table 3 |
| Results of individual studies | 20 | For each analysis in each study (e.g. unique combination of index test, reference standard, and positivity threshold) report 2x2 data (TP, FP, FN, TN) with estimates of diagnostic accuracy and confidence intervals, ideally with a forest or receiver operator characteristic (ROC) plot. | Results: Overall Diagnostic Accuracy (p. 6); Figure 2 |
| Synthesis of results | 21 | Describe test accuracy, including variability; if meta-analysis was done, include results and confidence intervals. | Results: Overall Diagnostic Accuracy (p. 6); pooled R@1 43.3% (95% CI 35.1-51.6; I^2=99.6%) |
| Additional analysis | 23 | Give results of additional analyses, if done (e.g., sensitivity or subgroup analyses, meta-regression; analysis of index test: failure rates, proportion of inconclusive results, adverse events). | Results: LLM Augmentation Strategy (p. 7); Benchmark Composition (pp. 6); Input Modality (p. 7); Figure 3 ; Table 2; Figure S1 (supplementary) |
| **DISCUSSION** | | |  |
| Summary of evidence | 24 | Summarize the main findings including the strength of evidence. | Discussion, paragraphs 1-2 (pp.8) |
| Limitations | 25 | Discuss limitations from included studies (e.g. risk of bias and concerns regarding applicability) and from the review process (e.g. incomplete retrieval of identified research). | Discussion, paragraph 4 (p. 9-10) -- limitations |
| Conclusions | 26 | Provide a general interpretation of the results in the context of other evidence. Discuss implications for future research and clinical practice (e.g. the intended use and clinical role of the index test). | Discussion, paragraphs 2-3 and 5 (pp. 9-10) -- implications for benchmarking, augmentation, and clinical validation |
| **FUNDING** | | |  |
| Funding | 27 | For the systematic review, describe the sources of funding and other support and the role of the funders. | Acknowledgements (p. 10); Abstract: Funding (p. 2) |

*Adapted From:* McInnes MDF, Moher D, Thombs BD, McGrath TA, Bossuyt PM, The PRISMA-DTA Group (2018). Preferred Reporting Items for a Systematic Review and Meta-analysis of Diagnostic Test Accuracy Studies: The PRISMA-DTA Statement. JAMA. 2018 Jan 23;319(4):388-396. doi: 10.1001/jama.2017.19163.

Page 2 of 2
