## Supplementary materials for "Diagnostic Accuracy of Large Language Models for Rare Diseases: A Systematic Review and Meta-Analysis"

**Web appendix to:**

**Diagnostic accuracy of large language models for rare diseases: a systematic review and meta-analysis**

### Search strategy

The search strategy combined two concept blocks across all databases. Block 1 (LLM terms) and Block 2 (rare disease terms) were combined with AND. The PubMed/MEDLINE strategy is shown below; equivalent strategies were adapted for Embase, Web of Science Core Collection, and Cochrane Library using database-specific syntax and controlled vocabulary where applicable. All searches covered January 2020 to February 2026 with no language restrictions.

#### *PubMed/MEDLINE*

**Block 1 — LLM and AI terms:**

> (“large language model”[tiab] OR “LLM”[tiab] OR “LLMs”[tiab] OR “GPT-4”[tiab] OR “GPT-3.5”[tiab] OR “ChatGPT”[tiab] OR “Gemini”[tiab] OR “LLaMA”[tiab] OR “PaLM”[tiab] OR “Med-PaLM”[tiab] OR “DeepSeek”[tiab] OR “Qwen”[tiab] OR “generative AI”[tiab] OR “foundation model”[tiab] OR “language model”[tiab] OR “retrieval-augmented generation”[tiab] OR “agentic”[tiab] OR “AI agent”[tiab] OR “multi-agent”[tiab] OR “Natural Language Processing”[MeSH])

**Block 2 — Rare disease terms:**

> (“rare disease”[tiab] OR “rare diseases”[tiab] OR “rare genetic disease”[tiab] OR “orphan disease”[tiab] OR “Mendelian disease”[tiab] OR “Mendelian disorder”[tiab] OR “monogenic disease”[tiab] OR “undiagnosed disease”[tiab] OR “undiagnosed patient”[tiab] OR “genetic syndrome”[tiab] OR “inborn errors”[tiab] OR “diagnostic odyssey”[tiab])

A supplementary MeSH sensitivity search added `"Rare Diseases"[MeSH]` to Block 2. This was applied to PubMed only; the Emtree equivalent inflated Embase results 4·6-fold, confirming that controlled vocabulary addition would compromise specificity in other databases.

#### *arXiv and medRxiv*

For preprint servers (arXiv sections: cs.CL, cs.AI, cs.LG, q-bio; medRxiv via Crossref API), nine keyword queries were used: “rare disease” AND “LLM”; “rare disease” AND “large language model”; “rare disease diagnosis” AND “agent”; “genetic diagnosis” AND “language model”; “phenotype” AND “LLM” AND “diagnosis”; “clinical genetics” AND “GPT”; “differential diagnosis” AND “rare” AND “AI”; “variant interpretation” AND “LLM”; “HPO” AND “LLM”. Results were deduplicated across queries and manually reviewed.

### Risk of bias assessment

Risk of bias was assessed using a modified QUADAS-3 instrument adapted for LLM-based diagnostic evaluation studies. The standard QUADAS-3 framework comprises four domains assessing participant selection, index test conduct, target condition definition, and flow and timing. We supplemented these with three AI-specific domains to address methodological concerns unique to LLM evaluations: data leakage, reproducibility, and evaluation fairness.

**Domain 1 — Participant selection** assessed whether the evaluation cohort was representative of the intended clinical population or whether case selection introduced spectrum bias (e.g., restricting to well-characterised diseases or enriching for diseases well-represented in LLM training data).

**Domain 2 — Index test conduct** evaluated whether the LLM system was applied as intended, including whether the prompt, model version, and configuration were consistent across all evaluation cases and whether the system had access to the reference standard diagnosis during evaluation.

**Domain 3 — Target condition definition** assessed whether the ground-truth diagnosis was established using an appropriate reference standard (e.g., molecular genetic confirmation) applied independently of the LLM output.

**Domain 4 — Flow and timing** evaluated whether all enrolled cases were included in the analysis and whether there was appropriate handling of inconclusive or failed LLM outputs.

**Domain 5 — Data leakage** (AI-specific) assessed the risk that evaluation cases or their diagnostic labels were present in the LLM's pretraining data, retrieval knowledge base, or fine-tuning dataset. This domain was rated high risk when evaluation benchmarks predated the LLM's training cutoff without temporal holdout validation, or when the system's knowledge base contained the benchmark's source data.

**Domain 6 — Reproducibility** (AI-specific) assessed whether sufficient methodological detail was provided to reproduce the evaluation, including exact model version, temperature and sampling parameters, prompt text, and code and data availability.

**Domain 7 — Evaluation fairness** (AI-specific) assessed whether all compared systems received equivalent prompting effort and whether the evaluation metric (R@1) was applied identically across systems, including consistency in disease name matching.

Each domain was rated as low risk (), high risk (+), or unclear (?). An overall judgment of high risk of bias was assigned if any domain was rated high. Assessments were performed independently by two reviewers, with disagreements resolved by discussion.

### Supplementary methods

#### *Phenopacket Store causal variant verification*

To characterise the information embedded in Phenopacket Store cases used by three included studies (Reese 2024, Chimirri 2025, MINT 2025), we programmatically inspected all phenopacket files in the repository (monarch-initiative/phenopacket-store, v0·1·26, GA4GH Phenopacket Schema v2·0·2). A total of 9,588 JSON files were inspected. Every case contained at least one pre-identified causal variant (mean 1·1 per case; 90·0% with one variant, 10·0% with two variants consistent with compound heterozygous inheritance). Of 10,547 variant entries, 94·4% included full VCF-format records (chromosome, position, reference and alternate alleles) and 99·9% carried an ACMG classification of pathogenic or likely pathogenic. This confirms that the Phenopacket Store embeds molecular diagnostic information alongside phenotypic descriptions, which is relevant when interpreting performance differences between studies that used HPO-only input versus those that incorporated the full phenopacket content.

#### *Orphanet prevalence mapping methodology*

To investigate whether benchmark disease composition contributed to observed performance heterogeneity, we mapped diseases from four commonly used evaluation benchmarks to Orphanet prevalence classifications using Orphanet Product 9 (epidemiology data, accessed March 2026).

For the Phenopacket Store (694 unique diseases), OMIM disease identifiers were extracted from 9,588 GA4GH phenopacket JSON files and cross-referenced to OrphaCode identifiers via the OMIM-to-OrphaCode mapping table (mapping rate: 49·1%). For RareBench (301 diseases), MAC Medline (301 diseases), and the Chinese Rare Disease List (66 diseases), disease names were mapped to Orphanet entries via programmatic search and manual curation (mapping rates: 88–91%).

Mapped diseases were classified according to Orphanet point prevalence categories: ultra-rare (<1 per 1,000,000), rare (1–9 per 1,000,000), or higher-prevalence (1 per 100,000). Diseases with multiple prevalence estimates were assigned their most specific geographic match. The resulting prevalence distributions showed substantial differences across benchmarks: the Phenopacket Store comprised 52·8% ultra-rare diseases, compared with 29·3% for RareBench, 30·1% for MAC Medline, and 1·7% for the Chinese Rare Disease List.

### Supplementary figures

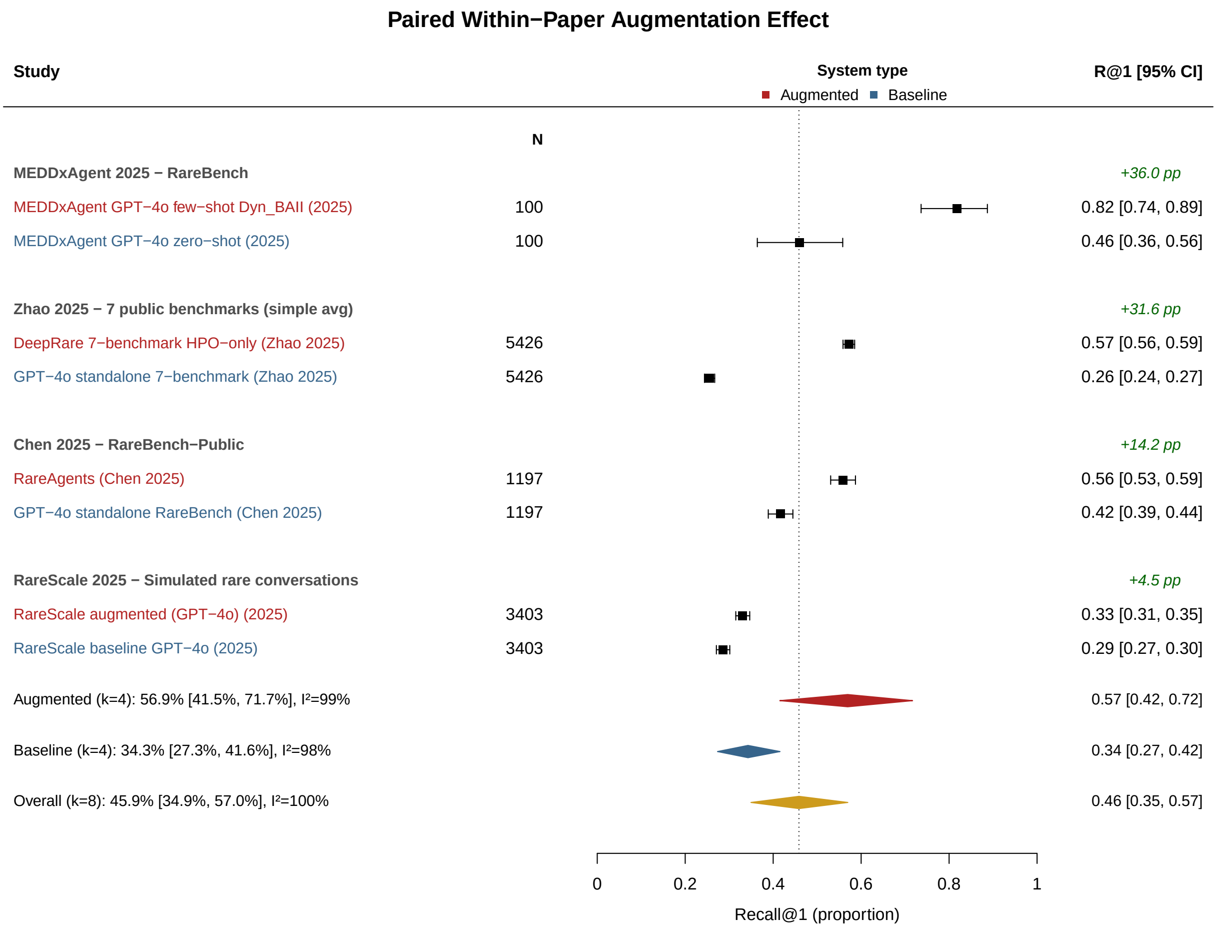

**Supplementary Figure S1:** Forest plot of paired within-study comparisons of augmented versus baseline LLM systems. Four studies provided matched comparisons on the same benchmark. In all four studies, augmented systems achieved higher R@1 than their baseline counterparts, with improvements ranging from 4·5 to 36 percentage points.

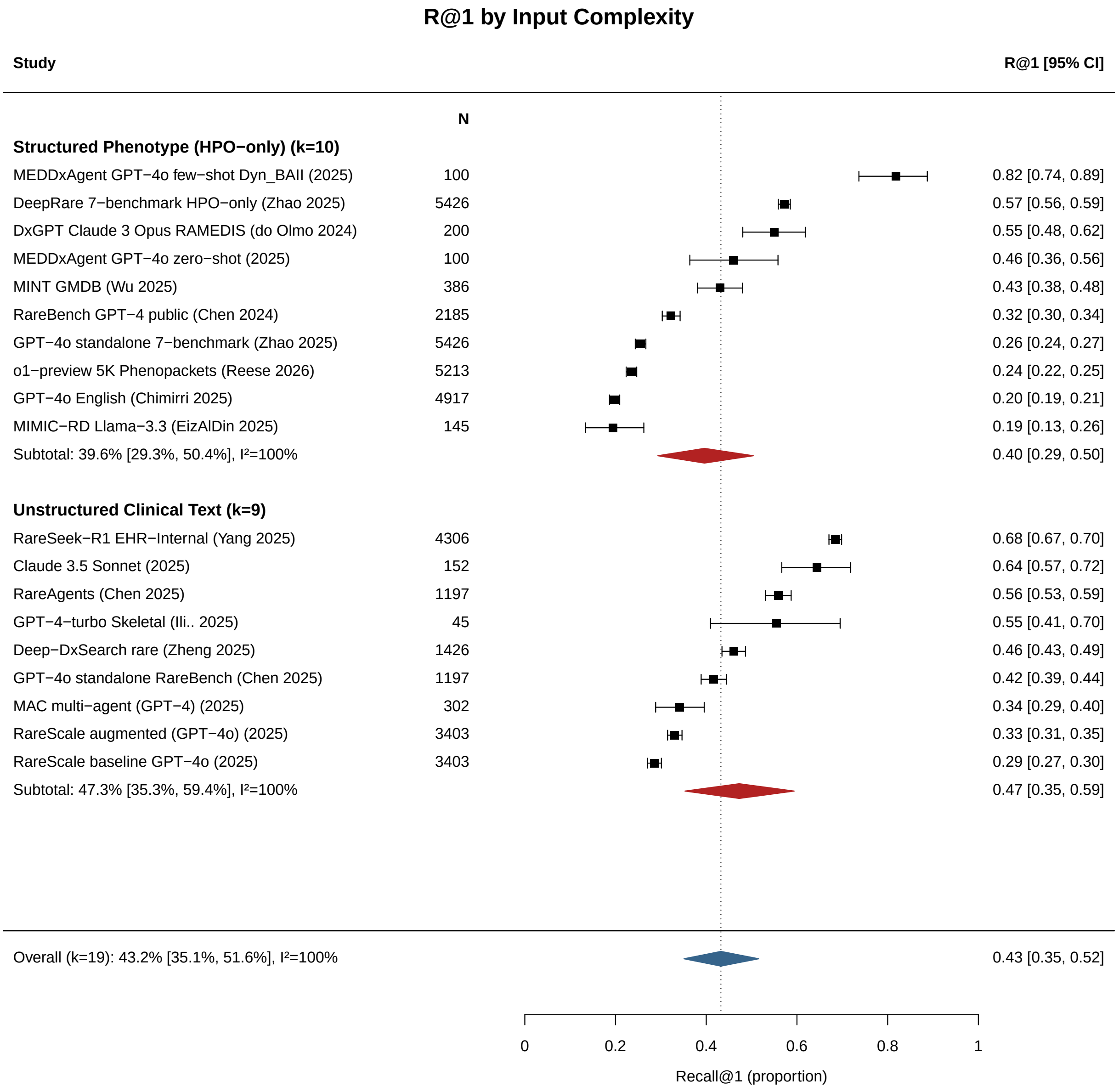

**Supplementary Figure S2:** Forest plot of subgroup analysis by input modality (HPO-only versus clinical text).

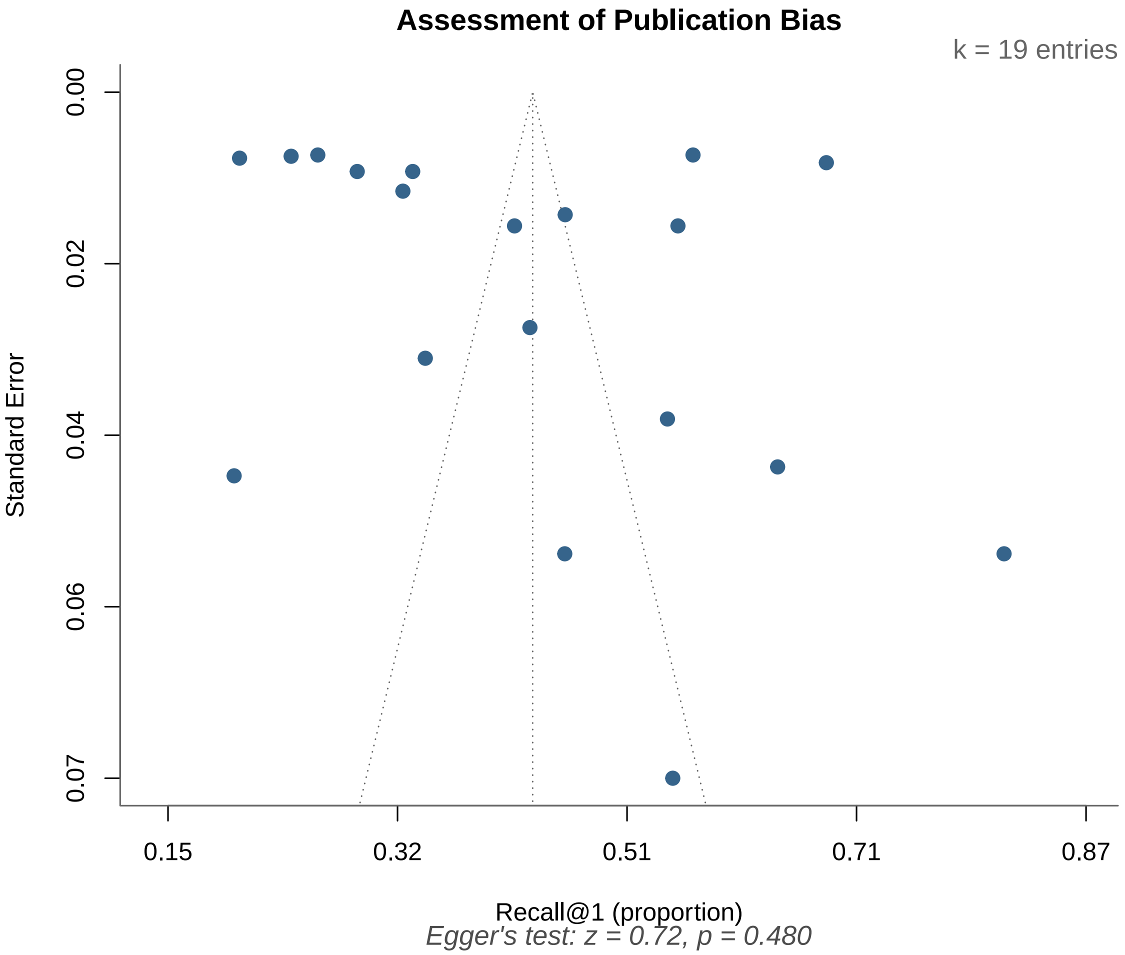

**Supplementary Figure S3:** Funnel plot for assessment of publication bias and small-study effects. The plot shows the distribution of study effect sizes against standard errors. Visual inspection does not suggest appreciable asymmetry.

### Supplementary table

Of the 15 included studies, 8 (53%) reported some form of prompt text. Three studies (20%) published complete verbatim prompt sets: DeepRare (14 numbered prompts in Supplementary Information Section 1.8), DxGPT (3 prompts in Appendix), and RareAgents (41 specialist system messages in code repository). Among the 8 augmented systems, 5 used explicit clinical role assignment, and 7 incorporated chain-of-thought or iterative reasoning. External knowledge was injected into prompts by 8 studies (53%). Two studies (MEDDxAgent, RareScale) provided explicit candidate disease lists. Four studies (DeepRare, MEDDxAgent, RareBench, Chimirri) included few-shot examples. Two studies (RareSeek, MINT) used fine-tuning rather than prompt engineering.

**Supplementary Table S1.** Prompting strategies and prompt component taxonomy for 15 included studies. Taxonomy columns: Role = clinical expert persona assigned; CoT = chain-of-thought or iterative reasoning; Out = output format specified; Cand = candidate disease list provided; Kn = external knowledge injected; Ex = few-shot examples. Y = present, N = absent. † = encoded implicitly via fine-tuning. ‡ = explicitly stated as absent.

| Study | Prompt | Role | CoT | Out | Cand | Kn | Ex |
|---|---|---|---|---|---|---|---|
| **Zhao (DeepRare)** | *14 prompts published verbatim in Nature Supplementary Information Section 1.8 (pp. 7–17). Key prompts:* | | | | | | |
| Prompt 1: Baseline diagnosis | "You are a specialist in the field of rare diseases. You will be provided and asked about a complicated clinical case; read it carefully and then provide a diverse and comprehensive differential diagnosis. Patient's {info_type}: {patient_info}. Enumerate the top 5 most likely diagnoses. Be precise, listing one diagnosis per line." | Y | N | Y | N | N | N |
| Prompt 2: Evaluation | "You are a specialist in the field of rare diseases. I will now give you five predicted diseases. Please identify the rank of the following gold-standard diagnosis. Please output the predicted rank; otherwise, output 'No'. Only output 'No' or '1-5' numbers." | Y | N | Y | N | N | N |
| Prompt 3: Rare/common discriminator | "You are a specialist in the field of rare diseases. Given the following HPO phenotypes: {patient_info}. Is this most likely associated with a rare disease or a common disease? Please reply with only one word: 'rare' or 'common'." | Y | N | Y | N | N | N |
| Prompt 4: Disease type classification | "You are a medical disease classifier specialising in categorising diseases into predefined categories." Classifies diseases into 14 body system categories (Blood/Heart/Circulation, Bones/Joints/Muscles, Brain/Nerves, etc.). Output: JSON format only with disease name and category array. | Y | N | Y | N | N | N |
| Prompt 5: Tentative diagnosis | "You are a specialist in the field of rare diseases. You have access to the following context: Online knowledge: {web_diagnosis}; LLM-generated diagnoses: {llm_response}; Diagnosis API results: {api_response}; Similar cases: {similar_cases}. Enumerate the top 5 most likely rare disease diagnoses with diagnostic reasoning and in-text citations." | Y | Y | Y | N | Y | N |
| Prompt 6: Gene analysis | "You are a specialist in the field of rare diseases. Here is a rare disease diagnosis case. Exomiser summary: {exomiser_summary}; HPO terms: {hpo_terms}; Preliminary diagnosis: {pheno_diagnosis}. Enumerate the top 5 most likely rare disease diagnoses, prioritising the Exomiser gene/variant results." | Y | Y | Y | N | Y | N |
| Prompt 7: Disease reflection | "Assume you are a doctor specialised in rare disease diagnosis. Evaluate whether the proposed diagnosis is correct for this patient. Begin with 'DIAGNOSIS ASSESSMENT: [Correct/Incorrect]'. Structure: 1. Patient summary; 2. Proposed diagnosis analysis; 3. References. Patient phenotype: {patient_info}. Medical literature: {disease_knowledge}." | Y | Y | Y | N | Y | N |
| Prompt 8: Final diagnosis | "Patient presentation: {patient_info}. Similar cases: {similar_cases}. Primary diagnosis results: {tentative_result}. Disease Reflection: {judgements}. Task: Enumerate the top 5 most likely rare disease diagnoses with 3–4 sentences of diagnostic reasoning and in-text citations." | N | Y | Y | N | Y | N |
| Prompt 9: HPO extraction | "Given a paragraph of patient information from discharge note, please extract the phenotype about this patient only. Check the HPO database. Only output the extracted phenotypes. Use the format: {'HPO': 'HP:0000000', 'Phenotype': 'description'}." | N | N | Y | N | N | N |
| Prompt 10: HPO normalisation | "You are a medical terminology translator specialising in rare diseases. Convert patient phenotype description into standardised HPO concept. Output JSON format only." (Includes 2 worked examples) | Y | N | Y | N | N | Y |
| Prompt 11: Knowledge summarisation | "Assume you are a doctor, please summarise these medical articles into a paragraph, only keep key message, mainly focus on the phenotype and related disease." | Y | N | Y | N | N | N |
| Prompt 12: Case comparison | "Assume you are a doctor experienced in rare disease diagnosis. Please judge if the two patient cases are likely to be the same disease based on the patient information. Only output 'Yes' or 'No'. Patient 1: {patient_info}. Patient 2: {retrieved_case}." | Y | N | Y | N | N | N |

*Table S1 continued*

| Study | Prompt | Role | CoT | Out | Cand | Kn | Ex |
|---|---|---|---|---|---|---|---|
| Prompt 13: Zero-shot LLM inference | "You are a specialist in the field of rare diseases. You will be provided and asked about a complicated clinical case; read it carefully and then provide a diverse and comprehensive differential diagnosis. Patient's {info_type}: {patient_info}. Enumerate the top 5 most likely diagnoses. Each diagnosis should be a rare disease. Use ** to tag the disease name." | Y | N | Y | N | N | N |
| Prompt 14: Rare disease judgment | "You are a specialist in the field of rare diseases. Given a disease name, determine whether it is a rare disease or a common disease. Output can only be 1/0." (Includes 4 worked examples) | Y | N | Y | N | N | Y |
| **Chen X (RareAgents), 2024** | *41 specialist system messages published in code repository. Example:* | | | | | | |
| MDT specialist | "As a [Specialist], you are dedicated to [specialist-specific clinical description]. Your expertise involves diagnosing and treating [domain]. Demonstrate proficiency in [domain assessments]. Communicate effectively with patients and families." (41 predefined departments) | Y | N | N | N | N | N |
| Diagnostic reasoning | Multi-round MDT discussion (up to R rounds). Phenomizer p-values, LIRICAL posteriors, and Phenobrain scores injected as tool outputs. Dynamic memory retrieves top-k=5 similar cases. | Y | Y | Y | N | Y | N |
| **Chen X (MAC), 2025** | Admin presents clinical case. 2–5 doctor agents provide diagnostic reasoning. Supervisor moderates up to 13 conversation rounds until consensus. Prompt text not published; AutoGen framework used. | Y | Y | Y | N | N | N |
| **Rose (MEDDx-Agent), 2025** | CoT prompting with dynamic few-shot examples selected via BGE-BASE-EN-v1.5 embeddings. "You may be provided a list of diagnosis options you can choose from. You must use this exact disease terminology." 102 unique diseases from RareBench subset. Wikipedia + PubMed knowledge retrieval. | N | Y | Y | Y | Y | Y |
| **Zheng (Deep-DxSearch), 2025** | System: "You are an AI assistant specialised in diagnosing rare diseases based on phenotypes. Your task is to analyse patient phenotypes, match them with similar cases, and identify the most likely diagnosis." Tools: Phenotype Matching (<match>), Disease Knowledge Search (<search>). Workflow: submit phenotypes → analyse matches → reason in <think> → query diseases → synthesise in <answer>. | Y | Y | Y | N | Y | N |
| **Yang (RareSeek), 2025** | 3-stage fine-tuning pipeline: (1) domain-specific instruction tuning, (2) CoT fine-tuning on 17,477 reasoning chains, (3) GraphRAG integration with Neo4j knowledge graph (635K ClinVar/HGMD variants, 7.5K OMIM entries, 4.3K Orphanet links, 16.7K HPO phenotypes). Generates up to 20 candidates normalised to Orphanet via MONDO API. | Y† | Y† | Y | N | Y† | N |
| **Wang (MINT), 2026** | Llama-3.2-3B-Instruct aligned via ORPO. Preference pairs from multimodal teacher (GestaltMML): "chosen" = teacher's top-ranked disease, "rejected" = lower-ranked. No explicit prompt engineering; 6.5K GMDB training samples. Input: HPO terms + demographics. | N† | N | Y | N | Y† | N |
| **Schumacher (RareScale), 2025** | *Three-stage prompt structure described but not published verbatim:* | | | | | | |
| Stage 1: Baseline DDx | "Generate 5 diagnoses without considering the rare candidate list." | N | N | Y | N | N | N |
| Stage 2: Candidate-augmented | "Consider the candidate list (but not required to use them; should discard if not appropriate)." Specialised 7B model provides 5-disease candidate list. | N | Y | Y | Y | Y | N |
| Stage 3: Final selection | "Select 5 most appropriate from both sets, including discarding all rare candidates if best." | N | Y | Y | Y | Y | N |
| **Chen X (RareBench), 2024** | | | | | | | |
| Zero-shot baseline | System: "You are a specialist in the field of rare diseases." Hit@k (k=1,3,10). CoT explored (32.3% zero-shot → 33.2% CoT). | Y | Y | Y | N | N | N |
| Dynamic few-shot | IC-weighted knowledge graph embeddings (HPO, OMIM, Orphanet; 17.2K phenotypes, 9.26K diseases) select m most similar training examples. m=3 boosted smaller models' R@1 by 108%. | Y | Y | Y | N | Y | Y |
| **Aldin et al., 2026** | "Given a patient's list of observed phenotypes, LLMs are asked to predict a list of the 10 most likely rare diseases in order of likelihood." Prompt text not published. | N | N | Y | N | N | N |

*Table S1 continued*

| Study | Prompt | Role | CoT | Out | Cand | Kn | Ex |
|---|---|---|---|---|---|---|---|
| **Chimirri et al., 2025** | phenopacket2prompt generates clinical vignette from GA4GH Phenopacket JSON (age, sex, observed/excluded HPO terms, onset). "We request the LLMs to return an ordered list of candidate diagnoses, giving an example output." | N | N | Y | N | N | Y |
| **Ilić et al., 2025** | Standardised clinical vignettes (demographics, prenatal history, skeletal findings, radiology, clinical course, biochemistry, family history). "Generate primary diagnosis + 2 differential diagnoses + confidence score (1–5)." Paper states: "No role-playing cues or guiding frameworks were supplied." | N‡ | N | Y | N | N | N |
| **Ao et al., 2025** | Clinical data excluding pathognomonic indicators (no genetic tests, biopsies, pathology). "Each model generated the top five most likely diagnoses, ranking them by probability." Prompt text not published (Letter). | N | N | Y | N | N | N |
| **Reese et al., 2026** | phenopacket2prompt converts Phenopacket JSON to clinical narrative (sex, age of onset, age at exam, observed/excluded HPO terms). "Return a differential diagnosis as a list of disease names." Zero-shot only. Prompts deposited on Zenodo. | N | N | Y | N | N | N |
| **Olmo (DxGPT), 2024** | "Behave like a hypothetical doctor who has to do a diagnosis for a patient. Give me a list of potential diseases with a short description. You have to indicate which symptoms the patient has in common with the proposed disease and which symptoms the patient does not have in common. Symptoms: {HPO_terms}" | Y | N | Y | N | N | N |

### PRISMA-DTA checklist

The PRISMA-DTA checklist is provided as a separate file (PRISMA+DTA+Checklist.docx) with locations where each item is reported in the manuscript.
